# How much of SARS-CoV-2 Infections is India detecting? A model-based estimation

**DOI:** 10.1101/2020.04.09.20059014

**Authors:** Srinivas Goli, K.S. James

## Abstract

**Background and Rationale:** Amid SARS-CoV-2 outbreak, the low number of infections for a population size of 1.38 billion is widely discussed, but with no definite answers.

**Methods:** We used the model proposed by Bommer and Vollmer to assess the quality of official case records. The infection fatality rates were taken from Verity et al (2020). Age distribution of the population for India and states are taken from the Census of India (2011). Reported number of deaths and SARS-CoV-2 confirmed cases from https://www.covid19india.org. The reported numbers of samples tests were collected from the reports of the Indian Council for Medical Research (ICMR).

**Results:** The findings suggest that India is detecting just 3.6% of the total number of infections with a huge variation across its states. Among 13 states which have more than 100 COVID-19 cases, the detection rate varies from 81.9% (of 410 estimated infections) in Kerala to 0.8% (of 35487 estimated infections) in Madhya Pradesh and 2.4% (of 7431 estimated infections) in Gujarat.

**Conclusion:** As the study reports a lower number of deaths and higher recovery rates in the states with a high detection rate, thus suggest that India must enhance its testing capacity and go for widespread testing. Late detection puts patients in greater need of mechanical ventilation and ICU care, which imposes greater costs on the health system. The country should also adopt population-level random testing to assess the prevalence of the infection.

## Introduction and rationale

The low numbers of SARS-CoV-2 cases in India given its population size presents a conundrum that is widely discussed, but with no definite answers. The low testing rate is often cited as a possible explanation for the low observed positive cases. There is considerable interest and need to understand the true rate of infection, but at the same time, it is difficult to obtain unless random tests are carried out at the population level. Since that may involve a long waiting period, what can be the possible predicted rates from available models across countries? In this context, we used the model proposed by Bommer and Vollmer to assess the quality of official case records.

## Methods

### Data

We have used data from multiple sources. The infection fatality rates were taken from Verity et al. (2020)^**1**^. Age distribution of the population for India and states are taken from the Census of India (2011)^**2**^. Reported number of deaths and SARS-CoV-2 confirmed cases from https://www.covid19india.org^**3**^. The reported numbers of samples tests were collected from the reports of the Indian Council for Medical Research (ICMR)^**4**^.

### Model

Researchers from the University of Gottingen used age-specific infection fatality rates from a study published in *The Lancet Infectious Diseases* journal^**5**,**1**^ to estimate the total number of infections. The details of Bommer and Vollmer (2020) model are presented elsewhere^**5**^. We have adjusted the age-specific infection fatality rates presented in the Lancet with the age distribution of the population of India and 13 states which have more than 100 confirmed cases as on 8^th^ April. The Lancet study reported perfect ascertainment between observed and estimated cases in the age group 50-59 years. We apply the infection fatality rate of this age group to all ages assuming infection attack rate is uniform for population across the age-groups in India. The number of infections is estimated as the number of deaths till April, 08, 2020 divided by the adjusted infection fatality rate. Based on the global evidence, we assumed that the mean duration between the onsets of symptoms to deaths is 14 days. Thus, it yields the total number of infections two weeks prior to April 8. The detection rate is estimated as the number of confirmed cases divided by the estimated number of infections. Further, by taking data from reports of the ICMR, we have also adjusted the detection rates with the sum of the difference in the rate of change in testing done on a daily basis to derive a more realistic figure of the total number of infections (confirmed + undetected infections) for India as of April 8.

## Results

Our estimates suggest that the total number of COVID-19 infections in India could be around 1, 59,939 instead of the 5,480 confirmed cases as on 8th April. As a result, the detection rate of infections for the country as a whole remains merely at 3.6%, below the world average of 6%. Based on the evidence from other countries, we assume that a major share (85%) of these cases may be those with milder symptoms, who may not have ended up in hospitals^**6**^. However, we found a huge variation within India. Among 13 states which have more than 100 COVID-19 cases, the detection rate varies from 81.9% (of 410 estimated infections) in Kerala to 0.8% (of 35487 estimated infections) in Madhya Pradesh and 2.4% (of 7431 estimated infections) in Gujarat. The detection rate in Maharashtra (the biggest COVID-19 infected state) is also very low with 1.8% (of 61,268 estimated infections). However, Rajasthan is moderately doing well in terms of the detection rate of infections by confirming 15.8% of total estimated infections. Even, economically and health systems wise better-off states such as Andhra Pradesh, Telangana, Tamil Nadu and Delhi are performing poorly in terms of detection rate; thus, experiencing more deaths than Rajasthan and Uttar Pradesh where more individuals were tested than estimated infections (Figure1 & Table 1).

**Figure 1:**
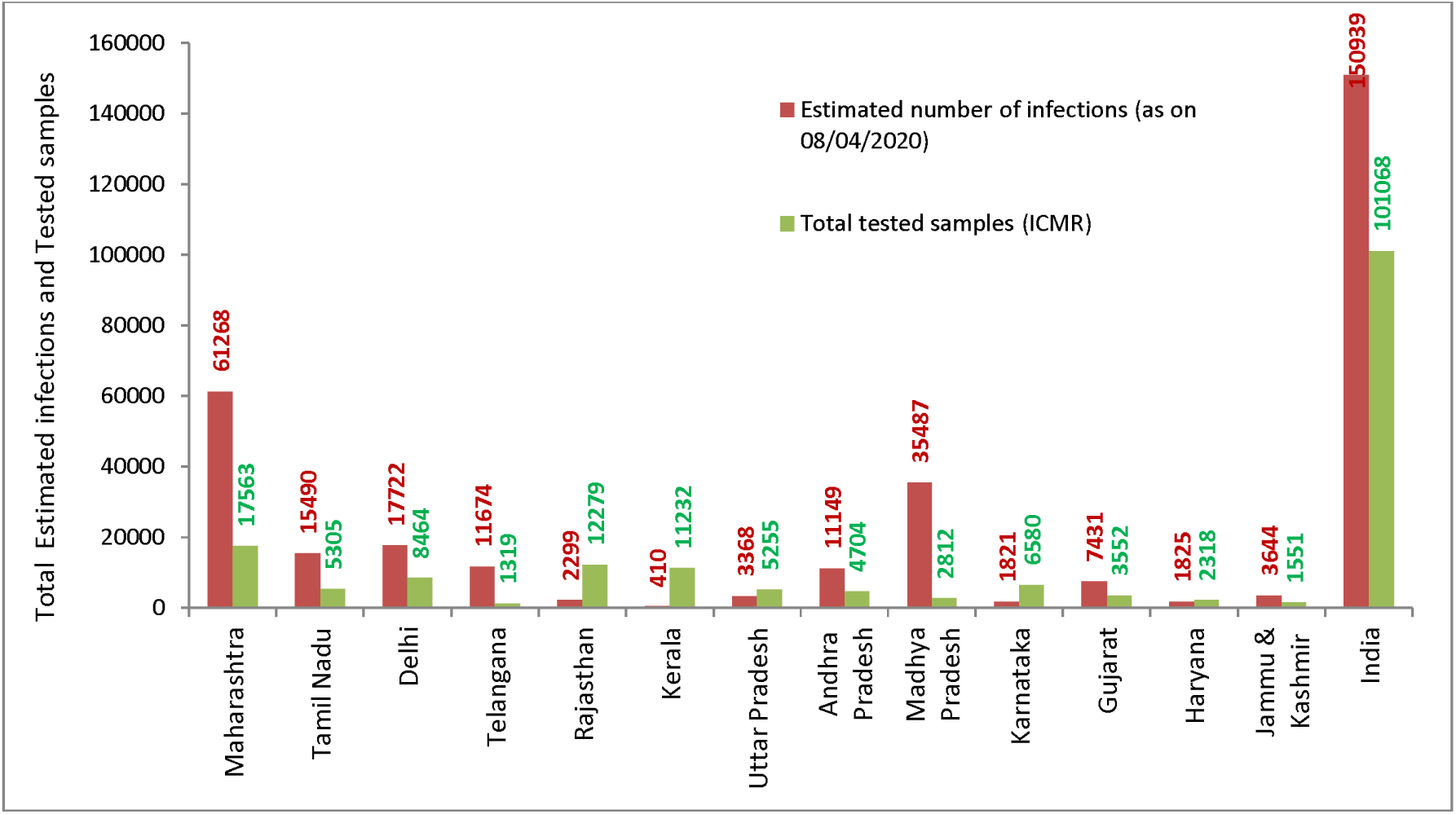
State-wise estimated total number of COVID-19 infections, Total tested samples and Detection rate in India as on 8^th^ April, 2020.

**Table 1:** State-wise estimated infection fatality rate, total number of infections and detection rate in India as of 8^th^ April, 2020.

| States | Infection fatality rate (%)<sup>1</sup> | Number of deaths (as on 08/04/2020) | Confirmed number of infections (as on 26/03/2020) | Estimated number of infections (as on 26/03/2020)<sup>2</sup> | Estimated detection rate (%)<sup>3</sup> | Number of confirmed cases (as on 8/04/2020) | Adjusted detection rate (%)<sup>♣</sup> | Estimated number of infections (as on 08/04/2020)<sup>4</sup> | Total tested samples (ICMR) | Date of update for total tested samples |
| --- | --- | --- | --- | --- | --- | --- | --- | --- | --- | --- |
| Col. | 1 | 2 | 3 | 4 | 5 | 6 | 7 | 8 | 9 | 10 |
| Maharashtra | 0.4359 | 66 | 130 | 15141 | 0.86 | 1078 | 1.76 | 61268 | 17563 | 6/4/2020 |
| Tamil Nadu | 0.5247 | 7 | 29 | 1334 | 2.17 | 690 | 4.45 | 15490 | 5305 | 7/4/2020 |
| Delhi | 0.3965 | 9 | 36 | 2270 | 1.59 | 576 | 3.25 | 17722 | 8464 | 6/4/2020 |
| Telangana | 0.432 | 11 | 43 | 2546 | 1.69 | 404 | 3.46 | 11674 | 1319 | 3/26/2020 |
| Rajasthan | 0.3583 | 2 | 43 | 558 | 7.70 | 363 | 15.79 | 2299 | 12279 | 5/4/2020 |
| Kerala | 0.5791 | 2 | 138 | 345 | 39.96 | 336 | 81.88 | 410 | 11232 | 7/4/2020 |
| Uttar Pradesh | 0.3356 | 3 | 43 | 894 | 4.81 | 332 | 9.86 | 3368 | 5255 | 5/4/2020 |
| Andhra Pradesh | 0.432 | 3 | 10 | 694 | 1.44 | 329 | 2.95 | 11149 | 4704 | 7/4/2020 |
| Madhya Pradesh | 0.3641 | 21 | 23 | 5768 | 0.40 | 290 | 0.82 | 35487 | 2812 | 5/4/2020 |
| Karnataka | 0.4409 | 5 | 55 | 1134 | 4.85 | 181 | 9.94 | 1821 | 6580 | 7/4/2020 |
| Gujarat | 0.4374 | 16 | 43 | 3658 | 1.18 | 179 | 2.41 | 7431 | 3552 | 7/4/2020 |
| Haryana | 0.3947 | 2 | 21 | 507 | 4.14 | 155 | 8.49 | 1825 | 2318 | 7/4/2020 |
| Jammu & Kashmir | 0.3587 | 3 | 14 | 836 | 1.67 | 125 | 3.43 | 3644 | 1551 | 5/4/2020 |
| **India** | **0.4053** | **167** | **730** | **41204** | **1.77** | **5480** | **3.63** | **150939** | **101068** | **6/4/2020** |

Note: †The method applied in this study is better predicts total number of infections for the states with more than 25 deaths. However, considering that there is a considerable chance of undercount of infections but also deaths, we modelled the estimates for all those states which greater than 100 cases on 8<sup>th</sup> April, 2020.
♣Detection rate estimated based on confirmed and estimated cases as on 26/03/2020 has been adjusted for the growth in the number of tests performed on a daily basis.
1. Col. 1. Is estimated using age-specific infection fatality rate from verity et al. (2020) after adjusting them to age-distribution of the population of India based on Census, 2011? 2. Col 4. Is estimated as $Co.2/(Col.1/100)$ 3. Col 5. Is estimated as $Co.3/(Col.4)*100$ 4. Col.8. Is estimated as $Co.6/(Col.7/100)$

Nearly 19.7% of all the COVID-19 infected cases are from Maharashtra alone, while its low detection rates pose a greater threat of COVID-19 related mortality in the state. Currently, the state contributes to 40% of the COVID-19-related deaths, and if the low detection rate continues, this number is likely to rise even further.

## Conclusion

Because states with a high detection rate (Kerala and Rajasthan) show lower death rates, we suggest that India must enhance its testing capacity and go for widespread testing. Late detection puts patients in greater need of mechanical ventilation and ICU care, which imposes greater costs on the health system. Early detection and avoidance of critical care for a majority of infected patients is an important way of avoiding overcrowding on the limited health care resources available in the country. The country should also adopt population-level random testing to assess the prevalence of the infection.

Also, there is a need to put out age-wise and co-morbidity characteristics of the patients who died from COVID-19 in the domain of epidemiologists, public health scientists and demographers to assess the relative risk of the infected population based on demographic characteristics. Such rapid assessments are critical to control the damage early and to ensure that resources are used efficiently.

## Data Availability

The data used in this study are public available and also presented in the paper. The relevant web-based links are already provided in the article.

https://www.covid19india.org

https://censusindia.gov.in/

https://icmr.nic.in/sites/default/files/whats_new/ICMR_testing_update_08April_9PM_IST.pdf

